# African Ancestry GWAS of Dementia in a Large Military Cohort Identifies Significant Risk Loci

**DOI:** 10.1101/2022.05.25.22275553

**Authors:** Richard Sherva, Rui Zhang, Nathan Sahelijo, Gyungah Jun, Tori Anglin, Catherine Chanfreau, Kelly Cho, Jennifer R. Fonda, J. Michael Gaziano, Kelly M. Harrington, Yuk-Lam Ho, William Kremen, Elizabeth Litkowski, Julie Lynch, Zoe Neale, Panos Roussos, David Marra, Jesse Mez, Mark W. Miller, David H. Salat, Debby Tsuang, Erika Wolf, Qing Zeng, Matthew S. Panizzon, Victoria C. Merritt, Lindsay A. Farrer, Richard L. Hauger, Mark W. Logue

**Affiliations:** National Center for PTSD, Behavioral Sciences Division, VA Boston Healthcare System, Boston, MA, 02130, USA; Boston University School of Medicine, Biomedical Genetics, Boston, MA, 02118, USA; VA Informatics and Computing Infrastructure (VINCI), Salt Lake City, USA; Massachusetts Veterans Epidemiology Research and Information Center (MAVERIC), VA Boston Healthcare System, Boston, MA, 02130; Division of Aging, Brigham & Women’s Hospital, Harvard Medical School, Boston, Massachusetts, USA; Department of Psychiatry, Boston University School of Medicine, Boston, MA, 02118 USA; Translational Research Center for TBI and Stress Disorders (TRACTS) and Geriatric Research, Educational and Clinical Center (GRECC), VA Boston Healthcare System, Boston, MA, 02130; Department of Psychiatry, Harvard Medical School, Boston, MA 02215; Department of Psychiatry, School of Medicine, University of California, San Diego, La Jolla, CA, USA; Center for Behavior Genetics of Aging, University of California, San Diego, La Jolla, CA, USA; Department of Medicine, University of Colorado Anschutz Medical Campus, Aurora, CO, USA; VA Eastern Colorado Healthcare System, Aurora, CO, USA; Center for Disease Neurogenomics, Departments of Psychiatry and Genetics and Genomic Science, Icahn School of Medicine at Mount Sinai, New York, NY 10029, USA; Center for Dementia Research, Nathan Kline Institute for Psychiatric Research, Orangeburg, NY 10962, USA; Mental Illness Research Education and Clinical Center (MIRECC), James J. Peters VA Medical Center, Bronx, New York, 10468, USA; Department of Neurology, Boston University School of Medicine, Boston, MA, USA; Boston University Alzheimer’s Disease Research Center, Boston University School of Medicine, Boston, MA, USA; Neuroimaging Research for Veterans Center, VA Boston Healthcare System, Boston, MA, USA; Geriatric Research, Education, and Clinical Center, VA Puget Sound Health Care System, Seattle, WA, USA; VA Washington DC Healthcare System, Washington, DC, USA; Center of Excellence for Stress and Mental Health, VA San Diego Healthcare System, San Diego, CA, 92161; VA San Diego Healthcare System, 3350 La Jolla Village Dr, San Diego, CA, 92161; Department of Biostatistics, Boston University School of Public Health, Boston, MA, 02118, USA; Department of Ophthalmology, Boston University School of Medicine, Boston, MA, USA; Department of Epidemiology, Boston University School of Public Health, Boston, MA, USA

## Abstract

We conducted the largest genome-wide association study (GWAS) of Alzheimer’s disease and related dementia (ADRD) in individuals of African-ancestry (AFR) to date using participants from the Million Veteran Program (MVP; 4,012 ADRD cases and 18,435 controls). A proxy GWAS based on survey-reported parental dementia (n=6,641 proxy cases, 45,970 controls) was also performed. The MVP AFR ADRD GWAS and proxy GWAS results were meta-analyzed and combined with the Alzheimer’s Disease Genetics Consortium’s (ADGC) AFR AD GWAS results. The MVP meta-analysis yielded genome-wide significant associations in or near *APOE, ROBO1*, and *RP11-340A13*.*2*. The MVP/ADGC meta-analysis yielded additional genome-wide significant variants near known risk genes *TREM2, CD2AP*, and *ABCA7*. We examined differences in expression of the implicated genes in a cohort of AD case and control brains. This study provides insight into dementia pathophysiology in historically understudied individuals of AFR and may help to address health disparities.

## Introduction

Late-onset Alzheimer’s disease (AD) is the most common form of dementia^1^. The prevalence of AD in the United States (US) differs by ancestry group, with a higher proportion of AD in African ancestry (AFR) population than European ancestry (EUR) population (see e.g. ^2; 3^). The difference in prevalence between ancestries is not fully understood^4^, and may be due to a combination of factors, including higher rates of health and cardiovascular conditions that are associated with dementia risk^5^ and societal factors which disadvantage AFR individuals in the US^6^.

Genetic factors also influence AD risk. The largest-effect genetic risk factor for AD is the *APOE* ε4 allele. Genome-wide association studies (GWASs) of EUR cohorts have increased our knowledge of the genetic architecture of AD beyond the *APOE* locus^7–9^. The most recent large-scale EUR GWAS of AD by Bellenguez et al.^10^, reported 75 AD risk loci in addition to *APOE*. Their study included 20,464 AD cases and 22,244 controls as well as n=46,828 “proxy” dementia cases in which case status was determined based on a reported history of AD or dementia in one or both parents. The sample sizes of non-EUR AD GWASs has been much smaller. Although there are consistent genes implicated in AD risk across different populations, the specific risk variants for a particular locus and the direction of effects for associated variants may differ by ancestry^11^. For example, the effect of the *APOE* ε4 locus in AFR cohorts is approximately half of that observed in EUR cohorts^1^. Thus, results obtained in one ancestral group may not generalize to other ancestries. Several AD GWASs have looked specifically at AFR cohorts^11–15^. The AFR AD GWAS by Reitz et al. (2013)^15^ included a cohort of 1,968 AD cases and 3,928 control participants, and, in addition to the *APOE* locus, identified a genome-wide significant association with a SNP (rs115550680, p=2.2×10^−9^) in *ABCA7*. A larger AFR AD GWAS was recently published by Kunkle et al. (2021)^12^ that included 2,784 screened AD cases and 5,222 controls from the Alzheimer’s Disease Genetics Consortium (ADGC) cohorts.

Although this was a small-scale study compared to the current EUR AD GWASs, 4 novel common loci were identified at the suggestive significance level (p<0.05×10^−7^), with variants in *EDEM1, ALCAM, GPC6*, and *VRK3*. A secondary analysis in Kunkle et al. that covaried for *APOE* genotype identified a genome-wide significant rare variant in an intergenic region on chromosome 15. In addition, they reported on gene expression data from EUR brain tissue showing associations between expression of *ALCAM, ARAP1, GPC6*, and *RBFOX1* and brain β-amyloid load. Other studies have implicated AFR-specific rare variants in the gene *AKAP9* in association with AD^16–18^. A trans-ethnic meta-analysis from 2017 identified additional AD risk variants in *PFDN1*/*HBEGF, USP6NL*/*ECHDC3, BZRAP1-AS1, NFIC*, and *TPBG*^11^. The work by Mez et. al. (2017)^14^ used a conditional liability model to identify risk variants in *COBL* and *SLC10A2*.

To identify AD-associated variants in AFR populations, we performed a GWAS in AFR participants of a large, national healthcare system biorepository, the Department of Veterans Affairs (VA) Million Veteran Program (MVP). These participants are of former US military service members who obtain their healthcare in the VA. MVP includes genetic data on MVP participants linked to their VA electronic medical record. While we are primarily interested in identifying AD risk variants, due to the limitations in the specificity associated with clinical diagnoses, we analyzed AD and related dementias (ADRD) cases and controls. We also conducted a “proxy dementia” GWAS of self-reported parental dementia, similar to the proxy GWAS included in recent EUR AD GWAS studies (e.g. ^10; 19^). This represents the largest GWAS of dementia in an AFR cohort, as well as the first use of proxy cases in AFR dementia GWAS meta-analyses. To increase power, we also performed a meta-analysis of the MVP-based GWAS with the results from the ADGC AFR AD GWAS and performed gene-based tests and pathway analysis in MVP as well as MVP+ADGC.

## Methods

### MVP Subjects and Diagnostic Classification Procedures

Subjects for this study included AFR MVP participants as determined by the genetically informed harmonized ancestry and race/ethnicity (HARE) method^20^. Consistent with other electronic medical record studies of dementia, ADRD cases were defined as participants with diagnosis codes for AD, non-specific dementia ^21^, or other related dementias diagnosis codes (i.e., vascular dementia, Lewy body dementia, frontotemporal dementia, and normal pressure hydrocephalus^22^), which are included in Supplementary Table 1. Because AD accounts for more than 50% of individuals with dementia^1^ and due to VA code usage^21^, it is likely that a large portion of MVP participants who were assigned ICD codes for non-specific dementia actually have AD. Further, we required two or more of these diagnosis codes and an age of onset (first diagnosis code) at or above age 60 to be included as a case. After exclusions for relatedness and other considerations (described in detail below), 4,012 participants met the ICD-code criteria for ADRD were included in the genetic analyses. A total of 18,435 individuals (representing approximately 4.5 times the number of ADRD cases) who were ages 65 and older without any dementia ICD codes, and who did not report a parental history of dementia in a health survey, were randomly selected as controls for the ADRD GWAS. The remainder were included as controls for the proxy GWAS described below.

The majority of MVP participants completed the “Baseline” survey which included questions regarding a wide variety of topics including employment, personality, psychiatric disorders, and family history of disease. For the proxy dementia GWAS, which excluded subjects used in the ADRD GWAS described above, participants who reported a history of “Alzheimer’s/ Other dementia” in their father or mother were classified as cases in the paternal or maternal proxy dementia GWASs respectively, and individuals with two affected parents contributed to both the maternal and paternal proxy analyses. Because the specific type of dementia is not confirmed or even recorded for parents of participants, we refer to this as a proxy dementia analysis and refer to any meta-analysis including these subjects as a meta-analysis of dementia rather than AD or ADRD to reflect the ambiguity in proxy case diagnosis. However, EUR GWAS results indicate a strong genetic correlation between proxy dementia GWAS and non-proxy AD case-control results in (r_g_=0.81)^19^. In order to limit the number of individuals with parents too young to be at risk for dementia, participants ages 45 and older without any dementia ICD codes (i.e. any codes from Supplementary Table 1) who were not included in the ADRD controls and did not report either parent as having dementia were included as proxy controls. Proxy cases were not screened on age. After applying these criteria, 4,385 maternal proxy cases, 2,256 paternal proxy cases, and a common proxy control cohort of 45,970 subjects remained for analysis.

### Genotype Data Generation and Processing

Genotype data processing and cleaning was performed by the MVP Bioinformatics core. The genotype data were generated using the MVP 1.0 custom Axiom array which assays 668,418 markers. Quality control (QC) included checks for sex concordance, advanced genotyping batch correction, and assessment for relatedness. The chip design and genotype cleaning pipeline have been described elsewhere^23^. The MVP Phase 4 genotype data release includes imputed genotype data for 62 million variants assessed for approximately 650,000 subjects. Imputation was performed using the African Genome Resources (AGR) panel from the Sanger Institute which includes the 1000 Genomes Phase 3 reference panel plus another 1,431 unrelated African-descent individuals to improve AFR imputation accuracy. Prior to imputation, SNPs with high missingness (20%), monomorphic, not in Hardy-Weinberg equilibrium, and that differed in frequency between batches were removed. Phasing was done with SHAPEIT4 v 4.1.3 and imputation was performed with MINIMAC4. One of each of a pair of related individuals, defined as a kinship coefficient of 0.09375 or higher, were removed from analysis using a scheme that prioritized a case over a control while the older control was selected if both were controls. When both members of a pair were ADRD cases, we selected a subject with AD-specific ICD codes or, in the absence of ICD codes, one individual was randomly selected. Principal components of ancestry were computed with FlashPCA2^24^, using only AFR individuals and a linkage-disequilibrium pruned set of 170,207 SNPs that excluded the major histocompatibility complex region of chromosome 6. *APOE* genotypes were determined using the “best guess” imputed genotypes (80% confidence threshold) for the rs7412 and rs429358 SNPs which were well imputed in the AFR cohort (r^2^= 0.87 and 0.99, respectively). The inclusion criteria for SNPs in SNP-and gene-based tests were set to that used in Kunkle et al. 2021^12^. Specifically, for the primary GWAS, SNPs with minor allele frequency (MAF)≥1% and imputation quality (r^2^)≥ 0.4 were included (N=16,589,632). For gene-based tests, SNPs with MAF≥0.1% and r^2^≥0.7 were included (N=20,289,645).

### Statistical Analysis

Association of each SNP with ADRD and proxy dementia was tested using logistic regression models implemented in PLINK 2.0. Firth logistic regression^25^ was applied when the standard regression model failed to converge. Models were adjusted for sex and the first ten ancestry principal components. GWAS for the maternal and paternal proxy AD/dementia outcomes were performed separately using a common set of controls, and the results from the maternal and paternal analyses were combined and meta-analyzed using the sample size weighted Z-score method as implemented in METAL including a correction for overlapping samples^26^. Results from the ADRD GWAS and proxy GWAS were then combined by weighted Z-score meta-analysis to generate the MVP AFR dementia GWAS results. As a sensitivity analysis, top hits were tested for association with a strict AD definition which included only cases with two or more AD ICD codes (see Supplementary Table 1 for codes, n=741) vs the non-demented controls described above. In addition, to match analyses performed in Kunkle et al,^12^ we also performed a GWAS adjusting for *APOE* ε4 allele dosage.

Gene-based tests, along with functional mapping and annotation, expression quantitative trait locus (eQTL) analyses, gene set enrichment tests, and canonical pathway analysis, were conducted using the FUMA web portal^27^ which uses the same methodology implemented in MAGMA^28^. Manhattan and regional plots were produced using FUMA. All presented genomic coordinates are according to GRCh37/hg19. Pairwise LD values were calculated based on 1000 genomes AFR population data using LDlink (https://ldlink.nci.nih.gov).

### Expression Profiling

We selected genes harboring variants with suggestive or genome-wide significant variants, as well as genes containing a SNP in LD (r^2^>0.6) with a suggestive SNP (including rare variants) identified by FUMA for further evaluation. As no comparably sized post-mortem AFR AD case-control cohorts exist, differential expression of these genes was assessed using RNA sequencing data generated from autopsied brains of 526 EUR AD cases and 456 controls from the Religious Orders Study/Rush Memory and Aging Project (ROS/MAP), the Mayo-Mount Sinai Brain Bank, the Framingham Heart Study, and the Boston University Alzheimer’s Disease Research Center Brain Bank^29^. As these were EUR cohorts, these data could not be used to validate specific associated loci as eQTLs, but were instead used to examine variations in expression profile of specific genes potentially relevant to dementia pathogenesis. The details of data generation, processing, and quality control were described elsewhere^29^. In brief, expression of each gene/isoform was compared between AD cases and controls using the LIMMA program including age of death, sex, and RNA integrity number as covariates. A Bonferroni correction was used to adjust for the number of genes examined.

## Results

The mean age at onset of ADRD cases was 73.8 years, which was similar to age at last exam for the controls (73.0 years). As typical for a cohort of older US Veterans, the proportion of women in the study was small; 3.0% of cases (n=120) and 5.9% of controls (n=1,090) were women.

### Association Findings in MVP

There was little evidence of genomic inflation in the ADRD GWAS (λ=1.01), the proxy GWAS (λ=1.006), or in the combined results from the ADRD and proxy GWAS (λ=1.11). Figure 1 shows the Manhattan and quantile-quantile (QQ) plots for the meta-analysis of the ADRD and proxy dementia GWASs.

**Figure 1:**
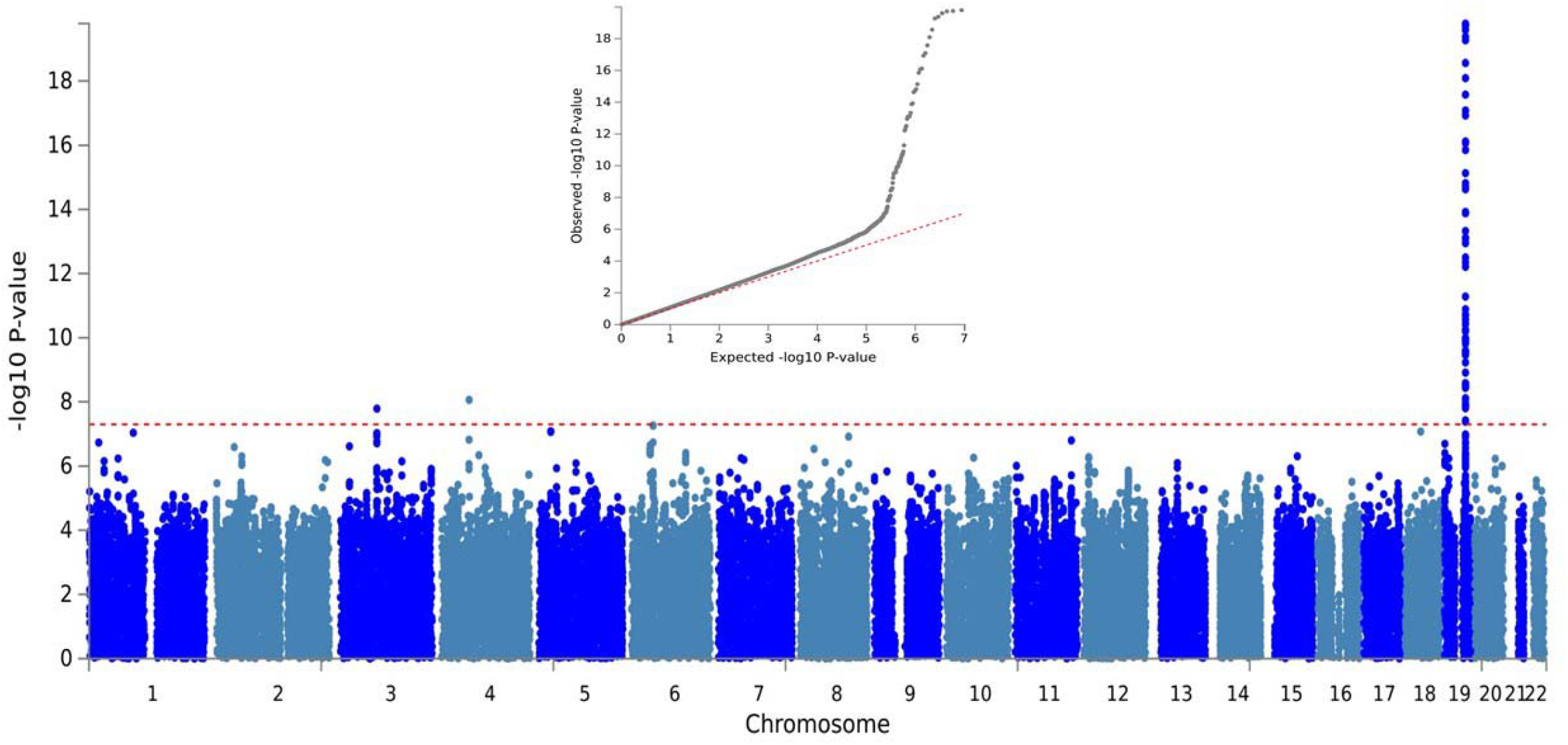
Manhattan and quantile-quantile plots for the meta-analysis of the Alzheimer’s disease and related dementias and proxy dementia genome-wide association studies in Million Veterans Program African Americans.

Genome-wide significant association was observed with *APOE* (Figure 2 and Supplementary Table 2). The odds ratios (ORs) of *APOE* ε2/ε4 and ε3/ε4 subjects were 1.45 (p=0.0001) and 1.44 (p=2.66×10^−18^), respectively, compared to ε3 homozygotes as a reference, which was approximately half of the effect size of APOE ε4/ε4 (OR=2.90) compared to the reference genotype (Supplementary Table 2). No genome-wide significant associations were observed for variants with MAF>1% with ADRD outside the *APOE* region.

**Figure 2:**
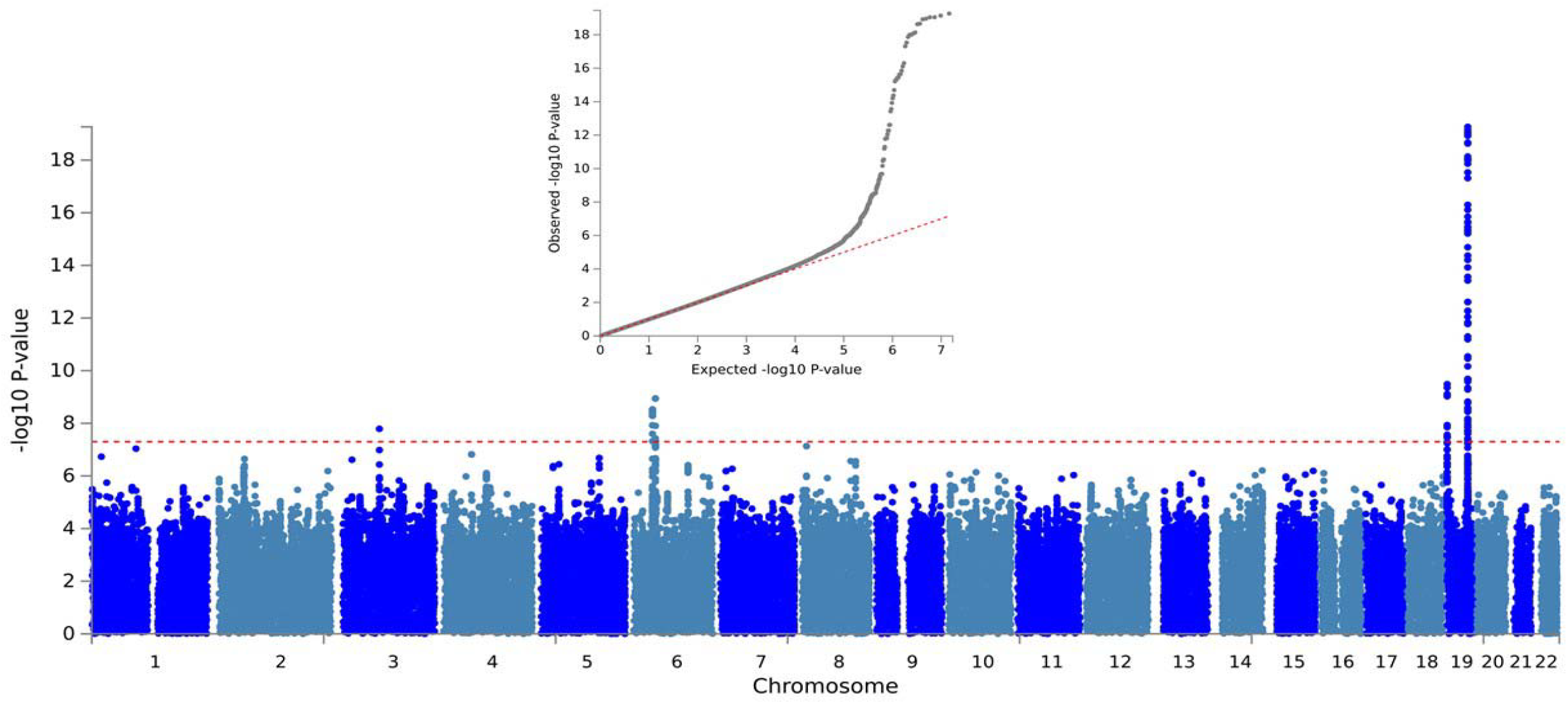
Manhattan and quantile-quantile plots for the meta-analysis of the Alzheimer’s disease and related dementias and proxy dementia genome-wide association studies in Million Veterans Program African Americans and the Alzheimer’s disease genome-wide association study in Alzheimer’s Disease Genetics Consortium African Americans. The red dashed line on the Manhattan plot represents the genome-wide significance threshold (5.0×10^−8^). Results from the *APOE* region on chromosome 19 are truncated at 1.0×10^−20^.

After meta-analysis with the MVP maternal-paternal proxy dementia results, two regions outside the *APOE* region showed genome-wide significant association: rs11919682 in the gene encoding roundabout guidance receptor 1 (*ROBO1*) on chromosome 3 (p=1.63×10^−8^) and rs148433063 in pseudogene *RP11-340A13*.*2* (aka *LINC02429*) on chromosome 4 (p=4.36×10^−9^; Supplementary Figure 1). There were also 15 suggestive (p<5.0×10^−7^) associations including SNPs near *CD2AP* (rs7738720), *TREM2* (rs2234253), and *ABCA7* (rs73505251) that were previously established as AD genes in EUR subjects (see Table 1 for details). Importantly, none of these were the same SNPs identified in recent GWASs in EUR cohorts^7–10^. Gene-based tests yielded one genome-wide significant association with *TREM2* (Bonferroni-corrected p=0.02), consistent with the significant *TREM2* gene-based association observed by Kunkle et al.^12^ Adjustment for *APOE* ε4 did not yield any genome-wide significant associations, but seven variants reached the suggestive level (Supplementary Table 3), including SNPs in or near *LHX6* (rs181518405, p=3.90×10^−7^), *CFAP46* (rs146777408, p=3.34×10^−7^), and *OCSTAMP* (rs202380, p=2.98×10^−7^).

**Table 1:** Common SNP associations (p<5.0×10^−7^) in the MVP ADRD+proxy dementia meta-analysis with corresponding results for AD in MVP and the ADGC

| SNP | Chr | Pos | Gene | A1/A2 | MVP<br>Meta<br>Z | MVP<br>Meta P | ADGC<br>B | ADGC P | MVP<br>AD B | MVP AD<br>P | MAF | $r^2$ |
| --- | --- | --- | --- | --- | --- | --- | --- | --- | --- | --- | --- | --- |
| rs10197243 | 2 | 55052493 | EML6 | T/C | 5.03 | 4.96E-07 | 0.09 | 1.60E-01 | 0.02 | 7.69E-01 | 0.15 | 0.89 |
| rs11919682 | 3 | 78770102 | ROBO1 | C/G | -5.65 | 1.63E-08 | NA | NA | -0.11 | 1.05E-01 | 0.27 | 0.85 |
| rs148433063 | 4 | 59886649 | RP11-340A13.2 | T/C | -5.75 | 8.65E-09 | 0.02 | 9.04E-01 | -0.64 | 2.99E-05 | 0.02 | 0.92 |
| rs28377689 | 4 | 80967671 | ANTXR2 | T/G | -5.04 | 4.59E-07 | 0.04 | 4.22E-01 | -0.15 | 1.91E-02 | 0.27 | 0.84 |
| rs74852218 | 5 | 25701222 | RNU6-374P | A/G | 5.36 | 8.20E-08 | 0.06 | 7.53E-01 | 0.69 | 2.48E-04 | 0.01 | 0.86 |
| rs2234253 | 6 | 41129105 | TREM2 | T/G | 5.18 | 2.23E-07 | NA | NA | 0.23 | 2.05E-03 | 0.13 | 1.00 |
| rs16894668 | 6 | 41388910 | AL136967.1 | T/C | 5.13 | 2.91E-07 | -0.01 | 9.18E-01 | 0.46 | 1.99E-03 | 0.02 | 0.99 |
| rs7738720 | 6 | 47395399 | AL355353.1 | T/C | -5.43 | 5.48E-08 | -0.20 | 5.51E-03 | -0.16 | 1.24E-01 | 0.10 | 0.76 |
| rs112395375 | 8 | 105698769 | RP11-127H5.1 | A/G | 5.29 | 1.22E-07 | 0.11 | 4.86E-01 | 0.76 | 1.81E-02 | 0.01 | 0.80 |
| rs509334 | 11 | 121277918 | RP11-142I2.1 | A/G | 5.24 | 1.59E-07 | 0.01 | 8.33E-01 | 0.16 | 3.54E-02 | 0.02 | 0.79 |
| rs116620371 | 18 | 34179992 | FHOD3 | A/G | -5.36 | 8.39E-08 | 0.11 | 6.07E-01 | -0.60 | 9.28E-02 | 0.01 | 0.92 |
| rs73505251 | 19 | 1068095 | ABCA7 | A/T | 5.20 | 2.03E-07 | 0.22 | 1.23E-04 | 0.17 | 2.19E-02 | 0.13 | 0.98 |

MAF
and $r^2$ represent the minor allele frequency and imputation quality in the MVP AFR cohort. A1/A2=effect/non-effect allele

As a sensitivity analysis, to examine whether the dementia-associated loci may be associated with AD specifically, we examined these SNPs in association with the strict AD phenotype. The *ROBO1* and *LINC02429* genome-wide significant SNPs and 9 of the 15 suggestive associations for ADRD were at least nominally associated with AD with the same effect direction (Table 1), noting, however, that the number of subjects with the strict AD phenotype was much smaller than those with the broader ADRD phenotype. Among the genome-wide significant and suggestive SNPs from the previous ADGC AFR GWAS implicated in either the *APOE* ε4-adjusted or unadjusted analysis, ten were evaluated in MVP, of which SNP rs9516245 in *GPC6* was nominally significant (p=0.03). The rare *IGFR1* SNP reported as genome-wide significant in the ADGC *APOE* ε4-adjusted analysis was not present in the MVP imputed results. Apart from *TREM2*, none of the other gene-based tests were nominally significant including those surpassing the p<1×10^−4^ threshold in the ADGC (Kunkle et al.) GWAS (i.e., *ARAP1, TRANK1, FABP2, LARP1B, TSRM, STARD10, SPHK1*, and *SERPINB13*).

### Dementia GWAS in Combined Datasets

Figure 2 shows the Manhattan and QQ plots for the full meta-analysis of the MVP and ADGC cohorts. After combining the GWAS results from the MVP and ADGC cohorts, variants in three known AD loci outside of the *APOE* region that were significant at the suggestive level in MVP alone emerged as genome-wide significant in the meta-analysis:*CD2AP* (rs7738720, p=1.14×10^−9^), a SNP upstream of *TREM2* (rs73427293, p=2.95×10^−9^) that is in high LD with rs2234253 (the peak variant in the MVP-alone analysis), and *ABCA7* (rs73505251, p=3.26×10^−10^; Figure 2, Table 2, Supplementary Figure 2). Of the two genome-wide significant associations observed in the MVP GWAS, rs148433063 on chromosome 4 was not significant in the ADGC dataset (p=0.9) or in the total sample (p=1.54×10^−7^). No result was available for the *ROBO1* SNP rs11919682 in the ADGC dataset. Notably, rs11919682 is a tri-allelic variant and may have been excluded from analysis in the ADGC dataset, although no copies of the rare alternate allele were present in the MVP imputed data. The second most significantly associated SNP in the *ROBO1* region, rs61053911 (p=9.36×10^−8^), is moderately correlated with the peak *ROBO1* SNP (r^2^=0.35) but was not significant in the ADGC dataset (p=0.66). SNPs in 17 additional regions were significant at the suggestive level including rs4607615 in *MSRA* that was nearly genome-wide significant (p=7.37×10^−8^). Results for three of these variants (rs567572378 in *KCNH8-EFHB*, rs192764155 in *RP11-506N2*.*1*, and rs2234253 near *TREM2*) were not available in the ADGC dataset. Seven rare variants (MAF<1%) showed suggestive evidence of association (Supplementary Table 4).

**Table 2:**
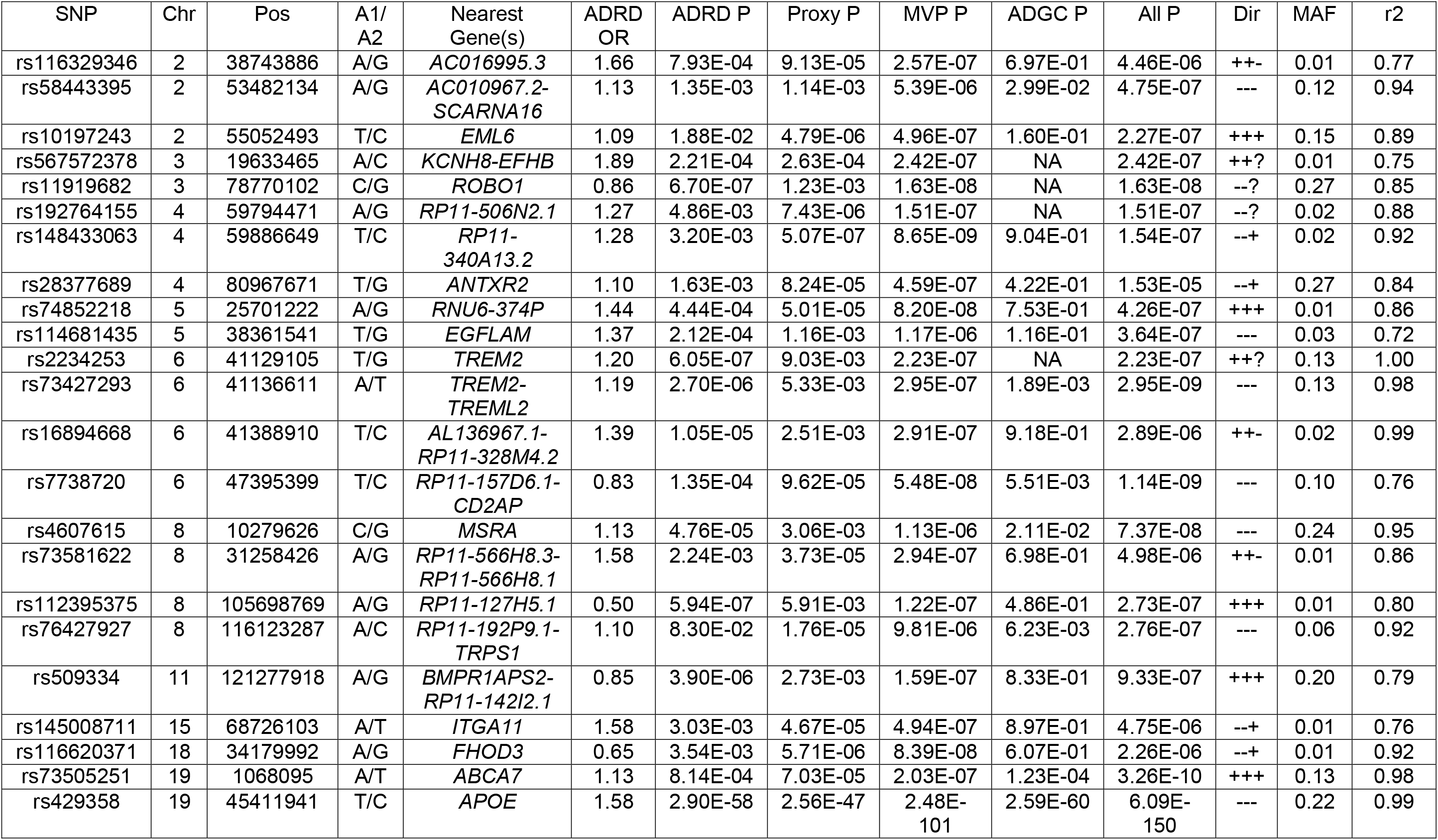
Common SNP associations (p<5.0×10^−7^) in the MVP ADRD+proxy dementia+ADGC AD meta-analysis

No additional genome-wide significant associations were found in the combined results from *APOE* ε4-adjusted models for ADRD in MVP and AD in ADGC, but there were two suggestive associations not observed in Model 1 including *RP11-144F15*.*1* (rs114572990, p=4.97×10^−7^) and *DNAI2* (rs9893381, p=4.89×10^−7^; Supplementary Table 5). Gene based tests for *TREM2* (Bonferroni corrected p=1.9×10^−5^) and *ABCA7* (Bonferroni corrected p=0.005) were significant in the total sample.

### Previously implicated AD variants in African Americans

Reitz et al.^15^ reported a genome-wide significant association with rs115550680 in *ABCA7* (p=2.2×10^−9^) but this SNP was not significant in a larger GWAS conducted by the ADGC (Kunkle et al.^12^) including the Reitz et al. sample (p=3.27×10^−5^). This SNP has again reached genome-wide significant (p=1.17×10^−8^) in our analysis combining the MVP and ADGC datasets. Of note, this SNP (rs115550680 in *ABCA7*) is not correlated with the top-ranked *ABCA7* SNP in the MVP study (rs73505251, r^2^=0.013), suggesting the possibility that these SNPs tag distinct functional variants. Neither the *ABCA7* frameshift deletion reported in Cukier et al.^30^ (rs142076058), nor the insertion/deletion polymorphism in the gene reported by Logue et al. (rs567222111)^16^ were nominally significant in the combined MVP sample (p>0.05). *TREM2* SNPs previously reported to be associated with AD in AFR studies, including a stop-gain mutation^31^, were significant in the combined MVP and ADGC datasets (rs7748513: p=6.06×10^−6^, rs2234256: p=4.635×10^−9^, rs2234258: p=5.48×10^−7^). All of these are in high LD with rs73427293, the peak *TREM2* SNP in the MVP+ADGC meta-analysis in terms of D’ (approximately 1.0), and moderate to strong LD in terms of r^2^ (0.14-0.99). Neither of the genome-wide significant SNPs in *COBL* and *SLC10A2* identified by Mez et al.^14^ were nominally significant in the full meta-analysis. Two rare AFR-specific non-synonymous *AKAP9* missense variants that were associated with AD in a smaller AFR sample^18^ were nominally associated with the combined MVP and ADGC datasets in a model adjusting for *APOE* ε4 (rs144662445, p=0.04; rs149979685, p=0.03), but neither was significant in the non-adjusted model. Among the tests of aggregated rare variants in genes ascribed to 74 distinct genome-wide significant loci reported in Bellenguez et al.^10^, only *TREM2, TREML2, CD2AP*, and *ABCA7* were significant in the combined MVP and ADGC datasets after Bonferroni correction (p<0.05).

### Functional Annotation

Because the most significant GWAS SNPs are often not the functional variants, we examined non-synonymous coding variants and eQTLs in LD with variants with genome-wide significant or suggestive evidence for association with AD. Excluding the *APOE* region, we identified two non-synonymous *ABCA7* variants (rs73505232, p=4.40×10^−10^, LD with peak SNP r^2^=0.87, and rs59851484, p=3.22×10^−8^, r^2^=0.66) that are in LD with rs73505251, the peak *ABCA7* SNP.

FUMA identified two candidate causal variants in LD with peak *TREM2* SNP: the previously-identified *TREM2* stop mutation and another nonsynonymous SNP (rs2234256 previously noted in^31^, p=4.64×10^−09^, LD with peak SNP r^2^=0.99 and rs2234253, p= 2.23×10^−7^, LD with peak SNP r^2^=0.99). Supplementary Table 6 shows the association results for non-synonymous variants in the region of the peak variants. FUMA additionally identified brain-tissue eQTLs in the region of the *ABCA7* and *MSRA* gene peaks according to GTEx version 8 (see Supplementary Table 7). A SNP in LD with the peak *ABCA7* SNP, rs34606911, is associated with *ABCA7* expression in the cerebellum and cerebellar hemisphere (p-value in dementia meta=9.16×10^−7^, r^2^=0.69 to peak *ABCA7* SNP). Several other SNPs in LD with the peak *ABCA7* SNP are brain eQTLs for flanking genes, including *GRIN3B, HMHA1, MUL1*, and *TMEM259*. FUMA also indicated six dementia-associated (p=2.03×10^−5^ to 9.83×10^−7^) *MSRA* eQTLs in LD with the peak *MSRA* SNP (r^2^ from 0.63 to 0.86) which are associated with MSRA expression in multiple regions of the brain, including the amygdala, cerebellum, nucleus accumbens, and the cortex. However, the GTEx database is comprised primarily of EUR samples, and hence many AFR-relevant eQTLs may have been missed.

### Pathway and Gene Set Enrichment Analyses

Gene set enrichment analyses in FUMA based on the gene-based tests indicated that a set including 8 genes involved in regulation of resting membrane potential was significantly enriched (Bonferroni corrected p=0.03). The enrichment was accounted for primarily by *TREM2* (gene-based p=1.00×10^−9^), although the gene-based tests for *PSEN1* (p=0.04), *KCNG2* (p=0.06), *KCNK6* (p=0.07), and *KCNK1* (p=0.07) also contributed to the enrichment signal. Five significant canonical pathways were identified and mostly accounted for by genes in the *APOE* region. These pathways were related to lipid metabolism, except for one involving immunoregulatory interactions between lymphoid and non-lymphoid cells for which the major contributors included *PVRL2, TREM2, TREML1*, and *TREML2* (Supplementary Figure 3). No significant enrichment was observed for gene sets or canonical pathways using results from the model adjusting for *APOE* ε4.

### Differential Expression

We examined differential expression of 30 genes with genome-wide significant or suggestively significant rare or common variants (Tables 2 and S4) or SNPs in LD with these variants in the post-mortem EUR AD case control cohort. Eighteen of 29 genes for which results were available showed significantly different expression between AD cases and controls, and nine remained significant after Bonferroni correction (p<0.05): *CDA, SH2D5, DCBLD1, EML6, GOPC, ABCA7, ROS1, TMCO4*, and *TREM2. CDA, ABCA7, TMCO4*, and *TREM2* were upregulated in AD cases and the other genes were downregulated (Table 3).

**Table 3:** Differential gene expression results for top genes in a meta-analysis of three European ancestry cohorts. P-values > 2.0E-04 not significant after multiple testing correction

| Gene | Chr | Meta P<sub>expression</sub> |
| --- | --- | --- |
| *CDA* | 1 | 1.60E-08 |
| *SH2D5* | 1 | 6.70E-07 |
| *DCBLD1* | 6 | 3.20E-06 |
| *EML6* | 2 | 6.60E-06 |
| *GOPC* | 6 | 7.50E-06 |
| *ABCA7* | 19 | 7.20E-05 |
| *ROS1* | 6 | 9.40E-05 |
| *TMCO4* | 1 | 1.30E-04 |
| *TREM2* | 6 | 2.00E-04 |
| *NUS1* | 6 | 2.20E-03 |
| *EIF4G3* | 1 | 5.30E-03 |
| *ARHGAP45* | 19 | 1.40E-02 |
| *CD2AP* | 6 | 1.70E-02 |
| *CNN2* | 19 | 2.80E-02 |
| *POLR2E* | 19 | 3.00E-02 |
| *RP11-127H5.1* | 8 | 3.10E-02 |
| *KIF17* | 1 | 3.40E-02 |
| *TREML1* | 6 | 3.90E-02 |

## Discussion

Increasing the representation of non-European populations in GWASs has been identified as a critical scientific and equity issue in genetic studies.^32; 33^ The differences in sample sizes between EUR and non-EUR studies to date could contribute to health disparities as a function of ancestry and the application of genetic information derived from EUR populations has reduced relevance and accuracy when applied to non-EUR populations^33; 34^. Moreover, the diversity gap in GWAS studies represents a missed opportunity because studies of traits in multiple populations have the opportunity to yield population-specific risk loci that can implicate new molecular pathways and potential therapeutic targets, as well as help to identify causal variants. This study, which utilized a large US Veteran cohort of AFR from the MVP project, therefore represents an important milestone in dementia genetics research.

This genetic study of ADRD, which included a sample that is more than twice the size of the previously largest AFR AD GWAS yielded several important findings relevant to AFR populations. First, using a large AFR cohort which included 4,012 ADRD cases, we examined the impact of the *APOE* AD risk locus genotypes. Whereas in the past, the literature had been inconsistent on whether AFR heterozygous carriers of the high risk ε4 allele were at increased AD risk (e.g. ^35,36^), here, it is clear that compared to the common ε3/ ε3 genotypes, those with either the ε3/ε4 and ε2/ε4 genotypes are indeed at increased ADRD risk (OR=1.44 and 1.45 respectively p<0.001). This risk was intermediate to ε4/ε4 risk of OR=2.66. However, we did not confirm the effect of the uncommon ε2 allele, which has been shown to be protective against AD in AFR and EUR. In our GWAS analyses, we confirmed the previously observed genome-wide significant AFR association with *APOE* and *ABCA7*. Additionally, we obtained the first genome-wide significant evidence for association in AFR cohorts with *TREM2* and *CD2AP*, known loci from EUR GWASs. We also found novel genome-wide significant associations with *ROBO1* and *RP11-340A13*.*2* loci.

The MVP GWAS meta-analysis also implicated several other genes whose impact on AD risk is less established. Not much is known about the function of *RP11-340A13*.*2* (aka *LINC02429*) that encodes a long intergenic non-protein coding RNA. However, there is strong biological rationale to suggest that *ROBO1* is an AD risk locus. *ROBO1* encodes the ROBO1 transmembrane receptor that binds SLIT proteins to regulate axon guidance and prevent axons from crossing the midline of the CNS^37; 38^. There is an extensive literature on the SLIT/ROBO signaling that mediates formation and maintenance of neural circuits in the hippocampus^39^.

Furthermore, SLIT proteins bind to both ROBO1 and APP receptors and their direct interaction triggers APP processing and ectodomain shedding, which can dysregulate APP-mediated axon pathfinding and contribute to AD pathophysiology^40; 41^. Additionally, *ROBO1* was recently implicated among the top genes in a study that examined single-variant and spatial clustering– based testing on rare variants in a whole-genome sequencing study of AD in a EUR population^42^, although this is the first time it has been identified as genome-wide significant. Because results for the associated *ROBO1* SNP were not available in the ADGC analysis and the association was not significant in the analysis of the strict AD phenotype (p=0.10), it is possible that this association may not be specific for AD but rather for dementia more broadly.

Several suggestive level associated SNPs in the MVP+ADGC meta-analysis are located in or near biologically plausible candidate genes including *MSRA, EML6, CDA*, and *GOPC*. Six SNPs that are in high LD with the peak *MSRA* SNP are significant eQTLs for *MSRA* in multiple brain tissues. MSRA reduces oxidative stress, and *MSRA* upregulation resulted in a marked reduction in aging phenotypes in transgenic mice^43^. Active in mitochondria, MSRA’s primary function is to reduce methionine sulfoxide, a potentially harmful reactive oxygen species produced during oxidative bursts, common during the neutrophil-and macrophage-mediated response to bacterial infection. We found that *EML6* was significantly downregulated in prefrontal cortex tissue from AD brains. *EML6* may modify the assembly dynamics of microtubules, making them longer and more dynamic, and is associated with sodium dependent hypertension^44^. In addition, we identified three genes in or near rare variants associated with ADRD at the suggestive level that are also differentially expressed in brain tissue between AD cases and controls: *CDA, SH2D5*, and *GOPC*. CDA catalyzes the hydrolytic deamination of cytidine and deoxycytidine to uridine and deoxyuridine and is a marker of monocyte to macrophage differentiation^45^. *SH2D5* encodes a mammalian-specific adaptor-like protein highly enriched in the brain and, based on the function of its homologue in mice, may modulate synaptic plasticity by regulating breakpoint cluster region protein Rac-GTP levels^46^. GOPC plays a role in intracellular protein trafficking and degradation and may affect AD pathogenesis by serving as a scaffold protein forming a complex between the CD46 receptor and Beclin 1^47^. Beclin 1 expression is downregulated in the entorhinal and frontal cortex during early AD, which would be expected to impair neuronal autophagy, dysregulate APP processing and increase &#x03B2;-amyloid deposition, and promote neurodegeneration and microgliosis^48^. In addition, GOPC suppresses complement attacks and inflammation^49^, is cleaved by the γ-secretase complex in response to microbial infection^50^, and impairment of these activities in AD could reduce microglial phagocytic capacity and amyloid-β clearance^51; 52^.

### Strengths and Limitations

This work represents the largest GWAS of AD and related dementias in individuals of AFR to date and is the first to focus on a veteran population. Veterans may be at increased risk for dementia due to higher rates of obesity-related health conditions, traumatic brain injury, and posttraumatic stress disorder^53; 54^. In addition, we obtained evidence that several of the most significantly associated genes are differentially expressed in the prefrontal cortex from AD cases and controls. Finally, SNPs in two novel loci (notably in *MSRA*) showed evidence for association in both the MVP and ADGC cohorts. However, several limitations to the study should be noted. First, because the sample size available for these analyses is small compared to AD GWAS in EUR GWAS studies, the opportunity for novel discovery is reduced and even some of the highly significant findings may not be robust. We did not observe corresponding association with the *ROBO1* and *RP11-340A13*.*2* in the ADGC data. This may be due to a lack of power, or from heterogeneity between the two cohorts. The MVP and ADGC cohorts differ in several important respects, including the proportion of males, the analysis phenotype, and the ascertainment scheme. It is also possible that these results represent false positives. However, the genome-wide significant associations observed with SNPs in AD loci previously established in other populations increases confidence that the findings with novel loci represent reliable associations. Second, because our findings were obtained from a sample including overlapping but non-identical phenotypes of AD, ADRD, and proxy dementia, the associations with novel loci may not be AD-specific. However, it is likely that most of these findings are AD-related given that AD is by far the most common form of dementia, the majority of the significant associations were observed with loci that were previously established in AD-specific cohorts, and ten of the 23 independent lead SNPs outside the *APOE* region were at least nominally associated with a strict AD diagnosis despite the substantially reduced sample size. Nonetheless, the novel genes identified in this study may not directly affect AD-related pathology but rather processes common to multiple types of dementia such as general cognitive function or memory. Finally, discordance of association findings between the MVP and ADGC cohorts in this study may reflect sex-specific influences noting that proportion of women differs widely between the MVP (approximately 10%) and the ADGC (approximately 60%) cohorts. Additional studies are needed to confirm and extend our findings in an enlarged AFR sample, as well as assess whether heterogeneity in the results between the MVP and ADGC AFR cohorts is due to the influence of unique environmental exposures for a largely male US Veteran cohort.

## Conclusions

The findings of this study build on the two known genome-wide significant common variant AD associations from AFR cohorts, *APOE* and possibly *ABCA7*, to include *TREM2, CD2AP, ROBO1*, and *RP11-340A13*.*2*. In addition, the top-ranked SNPs for *ABCA7, TREM2* and *CD2AP* differ between AFR and EUR studies suggesting the possibility of distinct causal variants in these populations. The loci and associations identified in this study represent a substantial increase in the knowledge of the genetic architecture of dementia risk in AFR populations. The insights they provide will pave the way for more accurate risk assessment for individuals of AFR. Moreover, the newly identified risk loci can contribute to a broader understanding of dementia pathogenesis which may provide targets for AD and dementia treatment.

## Data Availability

US Department of Veterans affairs restrictions do not allow the deposition of data into a public database. However, summary results will be availble on dbGAP once the study has been published. Additionally, any VA users with access to Million Veteran Program data will be able to confirm our data and results.

## Funding

This research is based on data from the Million Veteran Program, Office of Research and Development, Veterans Health Administration, and was supported by VA BLR&D award BX004192. This publication does not represent the views of the Department of Veteran Affairs or the United States Government.

## Disclosures

None of the authors have any conflicts of interest to disclose relating to this work.

## Author Contributions

Study design: ML, RS, Analysis: RS and RZ. Data Interpretation: RS, RZ, NS, GJ, MWL Phenotype Development in the MVP “Cognitive Decline and Dementia During Aging” Working Group: RS, RZ, TA, CC, KC, JF, JMG, KH, YH, WK, EL, JL, ZN, RP, DM, JM, MWM, DS, DT, EW QZ MP VM LF RH, and MWL Gene Expression Results: NS, GJ, Manuscript Drafting ML, RS, LF, GJ. Critical Editing and approval of the final manuscript: All authors.

## Data Availability

Because of VA rules of privacy, individual level data for MVP participants cannot be currently shared outside of MVP. However, the developed phenotypes and analysis code for this project have been deposited with MVP and can be made available to investigators within the VA with MVP access. Genome-wide summary statistics corresponding to the MVP AD, ADRD, proxy GWASs and the MVP meta-analysis will be deposited in dbGAP.

## Notes

### Competing Interest Statement

The authors have declared no competing interest.

### Author Declarations

US Department of Veterans Affairs Central Institutional Review Board protocol number 1613685-5

